# Evaluation of RC48-ADC in combination with PRaG regimen: An open-label, prospective, multicentre study assessing efficacy and safety for advanced refractory HER2-expressing solid tumors (PRaG3.0 Study Protocol)

**DOI:** 10.1101/2025.08.13.25333549

**Authors:** Meiling Xu, Yuehong Kong, Junjun Zhang, Rongzheng Chen, Pengfei Xing, Xiangrong Zhao, Shicheng Li, Yingying Xu, Liyuan Zhang

## Abstract

**Introduction:** Combining antibody-drug conjugates (ADCs) with radioimmunotherapy is a feasible and highly promising approach for treating HER2-positive patients, offering a potential paradigm for pan-cancer therapy. ADCs have demonstrated significant efficacy in cancers expressing human epidermal growth factor receptor 2 (HER2), independent of the tumor’s tissue of origin. Preclinical studies suggest that ADCs not only induce immunogenic cell death but also selectively enhance tumor radiosensitivity, providing a strong rationale for their integration with immunotherapy and radiotherapy. A combination regimen (PRaG) including hypofractionated radiotherapy (HFRT) alongside a PD-1 inhibitor and GM-CSF leverages HFRT to trigger tumor antigen release, GM-CSF to stimulate the proliferation and activation of antigen-presenting cells, and PD-1 inhibitors to relieve suppression of CD8+ T cells. The trial reported an objective response rate (ORR) of 16.7%, with three patients achieving complete remission. Building upon these findings, a next-generation regimen—disitamab vedotin (RC48) ADC combined with PRaG regimen (termed PRaG3.0)—may further amplify synergistic antitumor effects in HER2-expressing cancers. This precise combination therapy offers an innovative and exploratory approach for treating patients with HER2-positive or HER2-low tumors across various tissue types, potentially addressing the unique challenges associated with HER2 expression heterogeneity.

**Objective:** This study aims to investigate the effectiveness and safety of RC48-ADC combined with PRaG regimen for HER2-expressing advanced solid tumors.

**Methods and analysis:** This study is a prospective, single-arm, open-label, multi-center clinical trial designed as a basket study. Enrolled patients with confirmed HER2-expressing solid tumors (IHC 3+, 2+, or 1+) that had progressed after standard treatment or were intolerant to it were divided into three cohorts: pancreatic cancer, gynecological tumors, and others. Patients received RC48 (2 mg/kg) via intravenous injection on day 1, followed by subcutaneous GM-CSF at 200 µg from days 3 to 7 and interleukin-2 (IL-2) at 2 million IU from days 8 to 12. Radiotherapy was initiated on day 3, targeting one lesion with hypofractionated radiotherapy (2-3 fractions of 5 or 8 Gy). PD-1/PD-L1 antibodies were administered within one week after completing radiotherapy. Treatment was repeated every three weeks, and if there were no target lesions, radiotherapy could be discontinued, with RC48 given for at least six cycles. After achieving a complete tumor response, maintenance therapy with PD-1/PD-L1 antibodies continued until disease progression or intolerable toxicity occurred. The primary endpoint was the objective response rate (ORR).

**Ethics and dissemination:** The study protocol received approval from the Ethics Committee of the Second Affiliated Hospital of Soochow University (JD-LK-2022-121-02), as well as from all other participating hospitals. The clinical trial registration number is NCT05115500 and registration Date date is November 4, 2021.

## Introduction

Antibody-drug conjugates (ADCs) represent a breakthrough in oncology, offering a highly specific and effective therapeutic approach by combining the tumor-targeting precision of monoclonal antibodies with the potent cytotoxicity of chemotherapeutic agents^[1, 2]^. ADCs have demonstrated efficacy across a variety of HER2-expressing tumors, extending beyond the traditional tissue-of-origin paradigm^[3]^. HER2 expression, associated with aggressive tumor behavior and poor prognosis, is not limited to breast and gastric cancers but is also found in colorectal, ovarian, pancreatic, and lung cancers^[4, 5]^. Agents like trastuzumab deruxtecan and disitamab vedotin (RC48) have shown significant anti-tumor activity in these malignancies, including those with low HER2 expression, underscoring their versatility and broad applicability^[6-8]^.

Beyond their direct cytotoxic effects, ADCs can induce immunogenic cell death (ICD), leading to tumor antigen release and enhanced immune system activation. This dual mechanism positions ADCs as ideal candidates for combination therapies, particularly with immunotherapeutic approaches like immune checkpoint inhibitors^[9, 10]^. Their ability to target diverse tumor types and stimulate immune responses highlights ADCs as a cornerstone of pan-cancer therapy, paving the way for more effective and precise treatment strategies.

Radiotherapy, with its well-established safety profile, widespread clinical availability, and immune-activating potential, has gained attention as a valuable partner in combination therapies^[11, 12]^. The PRaG therapy—an innovative immunotherapy regimen—integrates hypofractionated radiotherapy (HFRT), PD-1/PD-L1 inhibitors, and granulocyte-macrophage colony-stimulating factor (GM-CSF) to create an in-situ vaccine effect, stimulating anti-tumor immunity and remodeling the tumor microenvironment. Grounded in the cancer-immunity cycle hypothesis, PRaG therapy addresses three critical stages: (1) HFRT releases tumor antigens, (2) GM-CSF activates antigen-presenting cells, and (3) PD-1 inhibitors restore CD8+ T-cell activity by counteracting inhibitory signals^[13]^. Widely implemented in China, PRaG therapy has shown notable clinical outcomes in treating advanced, refractory cancers, including esophageal, gastric, colorectal, pancreatic, lung, and breast malignancies, with demonstrated efficacy in refractory cases such as gastric cancer and ovarian cancer. A phase II trial reported an overall response rate (ORR) of 16.7% and a disease control rate of 46.3%, including complete remission in select patients^[14]^.

Disitamab vedotin (RC48), a novel humanized anti-HER2 ADC, incorporates monomethyl auristatin E (MMAE) as its cytotoxic payload and exhibits strong HER2 affinity and robust antibody-dependent cell-mediated cytotoxicity (ADCC)^[15-17]^. Beyond its cytotoxic effects, RC48 can induce ICD, promoting widespread release of tumor antigens and enhancing immunotherapy effectiveness by activating effector T cells. These properties make RC48 an attractive candidate for integration into multi-modal combination therapies^[18, 19]^.

The PRaG3.0 regimen, combining RC48-ADC with HFRT, PD-1/PD-L1 inhibitors, GM-CSF, and IL-2, represents a novel, synergistic treatment strategy targeting HER2-expressing cancers, including those with low HER2 expression. This approach leverages the complementary mechanisms of ADCs, radiotherapy, and immunotherapy to achieve enhanced anti-tumor effects. To evaluate the clinical potential of this paradigm, an exploratory phase II, open-label, multi-center, single-arm study was conducted, focusing on the efficacy and safety of PRaG3.0 in patients with advanced solid tumors exhibiting HER2 expression.

## Methods

### Objectives

The primary objective of this study is to investigate the effectiveness of RC48-ADC combined with radiotherapy, PD-1/PD-L1 inhibitor sequential granulocyte-macrophage colony-stimulating factor, and interleukin-2 for HER2-expressing advanced solid tumors. The secondary objective is to assess the safety and toxicity of this treatment. The study also aims to explore a panel of T lymphocyte subsets, tumor-associated cytotoxic T cells, activated cytotoxic T lymphocytes, activated memory T cells, monocytes, dendritic cells, interleukin-2, interleukin-4, interleukin-6, interleukin-10, interleukin-17A, tumor necrosis factor, interferon-γ.

### Study design and Sample calculation

The PRaG3.0 trial (NCT05115500) was a single-arm, open-label, multicentre, phase II study initiated by the Second Affiliated Hospital of Soochow University. The recruitment period for this study is from November 2021 to April 2026. The trial was designed as a basket study. Enrolled patients were divided into three cohorts that were pancreatic cancer, gynecological tumors, and the others. Simon 2 stage optimization design was adopted.

Pancreatic Cancer: The null hypothesis H0 posits an objective response rate (ORR) ≤0.05, whereas the alternative hypothesis H1 posits an ORR ≥0.2. With a one-sided α=0.05 and β=0.2, the calculated total sample size for a single-arm is 29. Stage 1 will enroll ten patients; at least one patient must respond positively in Stage 1 to proceed to Stage 2, which will enroll 19 patients. If the total number of positive responses after completing Stage 2 is greater than 4, the trial group is considered effective. If no patient responds positively in Stage 1, the study will be terminated.

Gynecological tumors: The null hypothesis H0 posits an ORR ≤0.05, whereas the alternative hypothesis H1posits an ORR ≥0.25. With a one-sided α=0.05 and β=0.2, the calculated total sample size for a single-arm is 17. Stage 1 will enroll nine patients; at least one patient must respond positively in Stage 1 to proceed to Stage 2, which will enroll eight patients. If the total number of positive responses after completing Stage 2 is greater than 3, the trial group is considered effective. If no patient responds positively in Stage 1, the study will be terminated.

Other Tumors: The null hypothesis H0 posits an ORR ≤0.05, whereas the alternative hypothesis H1 posits an ORR ≥0.2. With a one-sided α=0.05 and β=0.2, the calculated total sample size for a single-arm is 29. Stage 1 will enroll ten patients; at least one patient must respond positively in Stage 1 to proceed to Stage 2, which will enroll 19 patients. If the total number of positive responses after completing Stage 2 is greater than 4, the trial group is considered effective. If no patient responds positively in Stage 1, the study will be terminated.

### Inclusion criteria

The study inclusion criteria will be as follows:(1) Age ≥18 years; (2) Participants with advanced, confirmed HER2-expressing (IHC3+, 2+ or 1+) solid tumors that had progressed after standard treatment, or standard treatment intolerance were enrolled. Patients must have recurrent or metastatic late-stage solid malignant tumors with a confirmed pathological diagnosis or medical history. Furthermore, pathology must show HER-2 positivity (IHC 1+, IHC 2+, or 3+), and there must be no guideline-recommended standard treatment options, or the patient must be intolerant or explicitly refuse standard treatments due to personal preference. Additionally, patients should have identifiable measurable metastatic lesions; (3)No occurrences of congestive heart failure, unstable angina, or unstable arrhythmias in the past 6 months;(4) Patient’s performance status must be graded 0-3 according to the Eastern Cooperative Oncology Group (ECOG) scoring system, with a life expectancy assessment of ≥3 months; (5) No severe history of hematologic, cardiac, pulmonary, hepatic, renal abnormalities, or immunodeficiencies; (6) One week before enrollment, absolute T-lymphocyte count must be ≥0.5 times the lower limit of normal; neutrophils must be ≥2.0×10^9/L; AST and ALT must be ≤3.0 times the upper limit of normal (for liver cancer/liver metastatic cancer, ≤5.0 times the upper limit of normal); creatinine must be ≤3.0 times the upper limit of normal;(7) Patients must possess the capability to understand and voluntarily sign the written informed consent form.

### Exclusion criteria

Patients who meet any of the following criteria will be excluded: (1)Pregnant or breastfeeding women.(2)Patients with a history of other malignancies within the past five years, except cured skin cancer and cervical carcinoma in situ.(3)Patients with uncontrolled epilepsy, central nervous system diseases, or psychiatric disorders, which, in the investigator’s judgment, could significantly affect the ability to provide informed consent or interfere with medication adherence.(4)Clinically significant (active) heart diseases, such as symptomatic coronary artery disease, New York Heart Association (NYHA) Class II or higher congestive heart failure, severe arrhythmias requiring medication, or a history of myocardial infarction within the past 12 months.(5)Patients requiring immunosuppressive therapy due to organ transplantation.(6)Patients with known major active infections, or significant hematologic, renal, metabolic, gastrointestinal, endocrine dysfunctions, or other uncontrolled serious comorbidities as judged by the investigator.(7)Patients allergic to any components of the investigational drug.(8)Patients with a history of immunodeficiency, including those testing positive for HIV or suffering from other acquired or congenital immunodeficiency diseases, those with a history of organ transplantation, or those requiring long-term oral steroid therapy due to other immune-related diseases.(9)Patients with active acute or chronic tuberculosis (T-spot positive, chest X-ray showing suspicious tuberculosis lesions).(10)Other conditions that the investigator considers inappropriate for inclusion.

### Treatment scheme and modalities

Enrolled patients were treated using the PRaG 3.0 protocol, those received RC48-ADC(2.0 mg/kg d1, every 3 weeks), then HFRT (2-3 doses of 5-8Gy) was delivered for one metastatic lesion every other day, followed by GM-CSF(200 μg d3-7), sequential IL-2(2million IU d8-12), and PD-1/PD-L1 inhibitor was dosing within one week after completion of HFRT. After RC48-ADC combined with PD-1/PD-L1 inhibitor sequential GM-CSF and IL-2 for at least six cycles, then maintenance with PD-1/PD-L1 inhibitor was administered until disease progression or unacceptable toxicity. The specific treatment protocol is shown in Figure1.

**Figure1:**
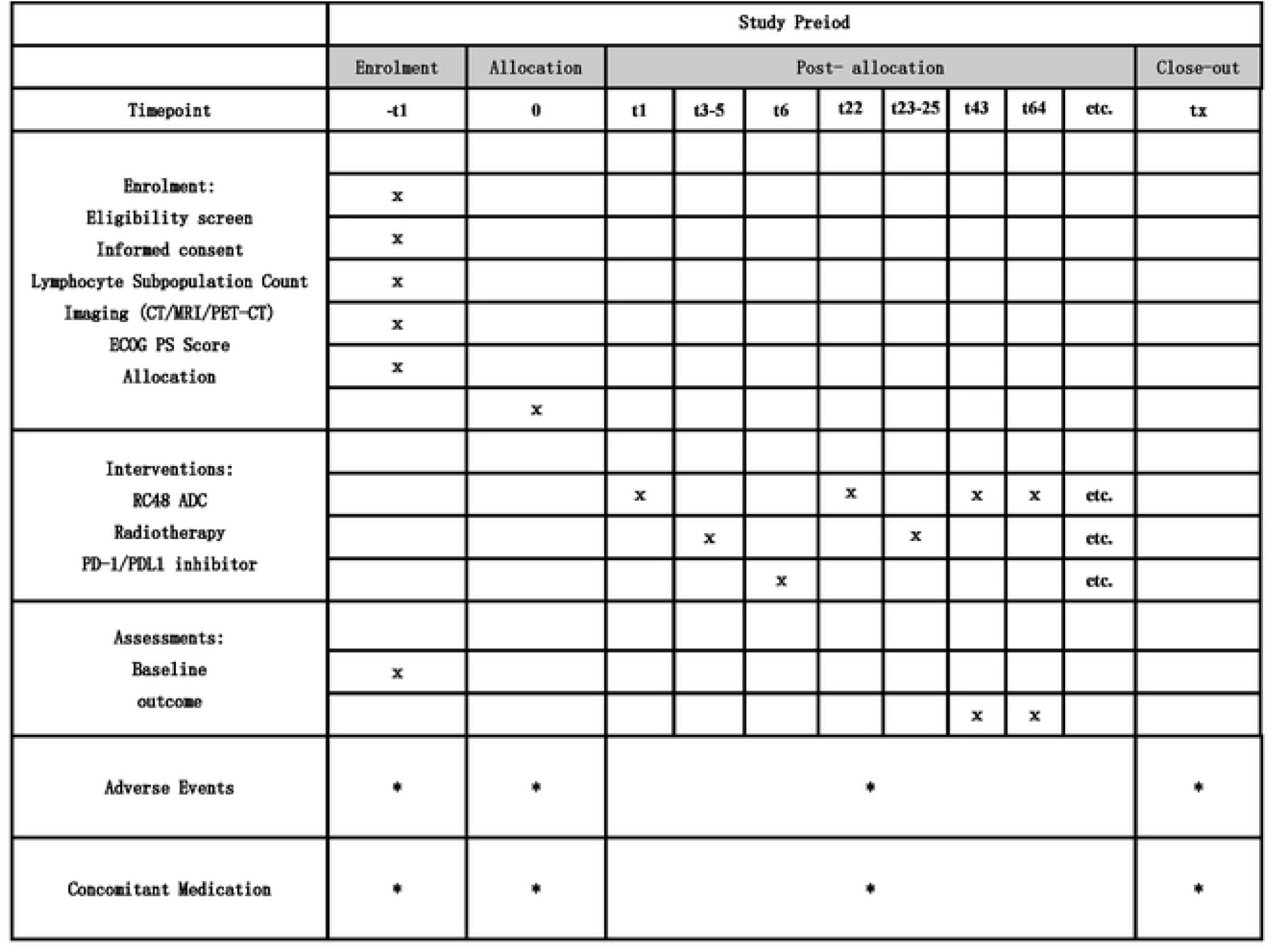
Treatment schedule of the PRaG3.0 therapy.

Patients received RC48 (2 mg/kg, IV) on day 1, GM-CSF (200 µg, SC) on days 3–7, and IL-2 (2 million IU, SC) on days 8–12. Radiotherapy (2–3 fractions of 5 or 8 Gy) began on day 3, followed by PD-1/PD-L1 antibodies within one week after radiotherapy.

### Objective endpoints and efficacy assessment

The primary endpoint was objective response rate (ORR), which was defined as the proportion of participants with partial (PR) or complete (CR) response in evaluable patients in accordance with the Response Evaluation Criteria in Solid Tumors (RECIST) 1.1 determined by investigators. Radiological assessments were performed on average every 6 weeks. Secondary objectives included safety, disease control rate (DCR), progression-free survival (PFS), and overall survival (OS). ORR was defined as the proportion of patients with complete response (CR) or partial response (PR). DCR was defined as the percentage of patients with CR, PR, or stable disease (SD) from enrollment. OS was calculated from the enrollment date to the date of death or last known alive. PFS was calculated from the enrollment date to disease progression, death, or censored at the last clinical follow-up. After the conclusion of treatment, all trial participants will undergo survival follow-up every 3 months until death, loss to follow-up, withdrawal of informed consent, or the sponsor decides to terminate the study. The nature, frequency, and severity of adverse events were assessed based on the Common Terminology Criteria for Adverse Events version 5.0 (CTCAE 5.0). Lymphocyte subset counts and cytokine analysis were examined as exploratory endpoints.

### Statistical analysis

Data analysis was conducted using SPSS 18.0 statistical software. Residuals were examined for normality using the Shapiro-Wilk test, with a significance level α>0.05. For variables that met the assumptions of normal distribution, a randomized block analysis of variance was employed; for those that did not, a non-parametric rank-sum test based on a randomized block design was utilized, with a significance level α<0.05. Comparisons were made for changes in the number of white blood cells, granulocytes, lymphocytes, and their subtypes, and cytokine levels before and after radiotherapy to determine if there were statistically significant differences. Survival time was considered in conjunction with these changes, using Cox regression analysis to assess the impact of changes in white blood cell counts, granulocyte counts, lymphocyte and its subtype counts, and cytokine levels on patient survival rates. Kaplan-Meier analysis was used to compare the survival rates between patients who experienced side effects and those who did not. The relationship between patient survival rates and associated cytokine levels was also analyzed using the Kaplan-Meier method.

### Patient and public involvement

Patients and/or the public were not involved in the design, conduct, reporting, or dissemination plans of this research.

### Ethics and dissemination

The study protocol has been approved by the Ethics Committee of the Second Affiliated Hospital of Soochow University and all other participating hospitals. It will be conducted in compliance with the Declaration of Helsinki. Informed consent will be obtained from each participant before the trial. The results of the PRaG3.0 study, regardless of the outcome, are intended to be published in a peer-reviewed international medical journal^[20, 21]^. The reporting of the trial’s findings will adhere strictly to the guidelines set forth in the Consolidated Standards of Reporting Trials (CONSORT) statement.

## Discussion

The results of this trial hold significant promise for advancing the treatment landscape of HER2-expressing advanced solid tumors, addressing key challenges associated with tumor heterogeneity and resistance to standard therapies. The PRaG3.0 regimen, integrating RC48-ADC with hypofractionated radiotherapy (HFRT), immune checkpoint inhibitors, GM-CSF, and IL-2, exemplifies the potential of combining targeted therapy, radiotherapy, and immunotherapy to achieve synergistic antitumor effects. RC48 inducing immunogenic cell death (ICD) not only enhances its direct cytotoxic potential but also primes the immune system for further activation, making it an ideal partner in this combination approach^[9, 15-19]^.

The preliminary results, which suggest a favorable objective response rate (ORR) across diverse tumor types^[14]^, highlight the versatility of this regimen in targeting HER2 expression regardless of tissue origin. This aligns with the broader trend of moving toward biomarker-driven, pan-cancer therapies^[22]^. Moreover, the dual role of HFRT in both local tumor control and immune activation underscores its importance as a central component of this strategy, particularly in facilitating antigen release and enhancing the efficacy of subsequent immune therapies^[11]^.

However, the study also brings to light several challenges and areas for further investigation. The safety profile of this multi-modal approach warrants close monitoring, given the potential for overlapping toxicities, particularly from ADC-related adverse events and immune-related complications. Additionally, the variability in HER2 expression levels (IHC 1+, 2+, 3+) across tumors may influence treatment efficacy, suggesting the need for further stratified analyses to identify optimal patient subgroups^[5, 23]^.

The exploratory nature of this trial provides a foundation for future research to refine the regimen, including adjustments in dosing schedules, biomarker-guided patient selection, and combination strategies with other novel agents. As HER2-targeted therapies evolve, this study underscores the importance of leveraging the unique mechanisms of ADCs like RC48 to enhance the effectiveness of radiotherapy and immunotherapy, potentially setting a new standard for personalized cancer treatment.

Future studies should focus on long-term outcomes such as overall survival (OS), progression-free survival (PFS), and quality of life, as well as the mechanisms underlying the observed immune modulation. The inclusion of correlative studies examining cytokine levels, immune cell populations, and tumor microenvironment changes will be critical to fully elucidate the biological basis of the observed clinical responses. With continued validation, the PRaG3.0 regimen could establish a transformative framework for treating HER2-positive and HER2-low tumors across multiple cancer types^[20, 21]^.

## Data Availability

yes?All XXX files are available from the XXX database.

## Author contributions

Study conception and design: MLX, YHK, JJZ, LYZ; Drafting of the trial protocol: MLX, LYZ; Critical review of the trial protocol for important intellectual content: RZC, PFX, XRZ, LYZ; Obtaining funding: LYZ; Coordinating investigator: SCL, YYX; Study implementation: MLX, YHK, JJZ, RZC, PFX, XRZ, SCL, YYX, LYZ; All authors read and approved the final manuscript.

## Ethics approval and consent to participate declaration

The trial was approved by the Ethics Committee of the Second Affiliated Hospital of Soochow University (JD-LK-2022-121-02). The study is registered in ClinicalTrails.gov (NCT0511550).

